# Palliative Care in Humanitarian Settings: An International Survey on Perceived Importance and Readiness among Health Emergency Response Unit Delegates

**DOI:** 10.64898/2026.03.12.26348178

**Authors:** Hanna Kaade, Susann May, Matthew Allsop, Marcel-Alexander Kamp, Martin Heinze, Felix Mühlensiepen

## Abstract

**Background:** Palliative care is recognised as an essential component of humanitarian health response, yet its delivery in field operations remains limited. This study assessed perceived importance, readiness to deliver, training needs, and operational barriers among Red Cross and Red Crescent Health Emergency Response Unit (ERU) delegates.

**Methods:** A cross-sectional, web-based survey (LimeSurvey) was conducted among health professionals with ERU deployment experience between 1 October and 31 December 2024. The questionnaire captured demographics, background, preparedness, barriers, and training preferences. Descriptive statistics summarised categorical variables, and free-text responses underwent content analysis by two reviewers. Quantitative and qualitative data were analysed separately and integrated during interpretation.

**Results:** Of 173 responses, 114 met inclusion criteria ((11) ≥50% completion). Half (50.9%) had over ten years of humanitarian experience. Most (71.1%) considered palliative care extremely important, yet only 49.1% reported providing it, usually limited to pain relief; 25.4% reported none. Barriers included insufficient time or resources (56.1%), lack of training (49.1%), absent policies (48.2%), cultural barriers (47.4%), limited knowledge (36.8%), and restricted opioid access (28.1%). Among prescribers, 85.1% felt comfortable prescribing opioids, but stockouts (54.2%) and regulations (44.9%) constrained use. Most delegates (75.4%) had delivered bad news without structured communication training. Overall, 82.7% reported no palliative care training, although 91.4% endorsed dedicated blended learning combining online and practical components.

**Conclusions:** ERU delegates view palliative care as essential yet under-implemented. Integrating core competencies, standard protocols, and medicine access pathways—supported by competency-based training—could strengthen humanitarian readiness and support the integration of palliative care within EMT standards.

## Introduction

The 2010 Haiti earthquake, which claimed an officially reported 316,000 lives and displaced more than 1.5 million people, revealed profound shortcomings in the global medical response. External aid was widely characterised by fragmented coordination, supply gaps, and uneven quality of care for survivors.^1–3^ Beyond the tragic human toll, Haiti became a watershed for global health response governance: the event demonstrated that ad hoc deployments, however well-intentioned, are insufficient in the absence of shared standards, interoperable systems, and accountability mechanisms.^1–3^

In the years that followed, the World Health Organization (WHO) developed a common language and set of expectations through *Emergency Medical Teams: Minimum Standards for Clinical Care in Disaster Situations* (the “Blue Book”), first issued in 2013 and updated in 2021.□ The framework clarifies Emergency Medical Team (EMT) typology (i.e., Type 1 outpatient—fixed or mobile; Type 2 inpatient surgical; Type 3 referral hospital) and details cross-cutting requirements for clinical governance, logistics, supply chains, data, and coordination, with the aim of ensuring consistent, high-quality care under extreme pressure.□ Crucially, the 2021 update explicitly includes palliative care —the management of pain, symptoms, and psychosocial distress — as a required clinical capability, reinforced by WHO’s operational *Framework for Palliative Care in Humanitarian Emergencies*.□,□ This reframing recognises that alleviating suffering is part of “doing everything possible,” not an admission of therapeutic failure.

Within this global framework, the Red Cross and Red Crescent Movement’s Health Emergency Response Units (ERUs) represent standardised, internationally deployable medical teams that predate but closely align with WHO’s EMT principles.□,□ Operated by the International Federation of Red Cross and Red Crescent Societies (IFRC) in collaboration with participating National Societies, ERUs can be mobilised within 24–48 hours to deliver both clinical and public health interventions when national capacities are overstretched. These multidisciplinary teams—comprising specialists in emergency medicine, surgery, obstetrics, paediatrics, anaesthesia, and nursing, supported by logisticians and public health experts—are capable of establishing fully functional field hospitals or clinics in austere environments. Their clinical services include fixed and mobile emergency clinics (comparable to WHO EMT Type 1), field hospitals with operating theatres, and maternal and newborn health units staffed by midwives and obstetric specialists (aligned with WHO EMT Type 2). Complementary public health services encompass community-based surveillance for disease outbreaks, case management of malnutrition and cholera, safe and dignified burials, and psychosocial support.□

These Health ERUs have demonstrated strong capacities in trauma and acute care, infectious disease control, and maternal and child health. However, palliative care expertise is rarely embedded, and frontline clinicians often report feeling underprepared to deliver comfort-focused care for patients with advanced or life-limiting illness.□,□ A growing body of literature documents a persistent implementation gap: while there is broad consensus that palliative care is ethically and clinically necessary during crises, its integration into humanitarian health responses remains limited.□–^12^ Barriers include insufficient training, restricted access to essential medicines (especially opioids), operational constraints, and ethical tensions in resource allocation.^13^ Clinicians frequently describe moral distress when balancing urgent, life-saving interventions with the time-intensive communication, symptom relief, and family support that palliative care requires—needs that do not disappear in emergencies but often become more acute.

Against this backdrop, this study explores how Health ERU delegates perceive and implement palliative care in humanitarian contexts, identifies priority training needs, and describes practical challenges encountered when striving to relieve suffering in humanitarian field settings. The findings aim to inform integration of palliative care competencies, resources, and operational guidance within existing WHO EMT and IFRC ERU preparedness frameworks.

This study aimed to assess the readiness of the Red Cross and Red Crescent Health Emergency Response Unit (ERU) delegates to provide palliative care in humanitarian missions, focusing on their perceived importance of palliative care, current knowledge levels, and practical preparedness. Specifically, it sought to: (i) assess perceptions of palliative care importance in emergency settings; (ii) evaluate self-rated knowledge and preparedness in pain relief, symptom management, and communication of bad news; (iii) explore prior and desired training in palliative care; (iv) examine availability of resources and infrastructure to support delivery; and (v) identify operational, logistical, and contextual challenges to implementation.

## Methods

### Study design and purpose

Given the increasing emphasis on integrating palliative care into humanitarian operations, this descriptive cross-sectional survey was conducted to assess the experiences and perceived challenges of integrating palliative care among Red Cross and Red Crescent Health ERU delegates. The study aimed to evaluate delegates’ readiness to deliver palliative care, understand its perceived importance, identify training needs, and document field-level barriers to implementation.

### Setting

Data were collected from 1 October to 31 December 2024 using an anonymous, self-administered online survey in English, hosted on the LimeSurvey platform. One reminder email was circulated midway through data collection. This was a *closed survey*, accessible only via a secure LimeSurvey link shared through internal IFRC and ICRC mailing lists (clinical delegate pools) and designated National Society focal points maintaining Health ERU delegate rosters. No public advertisements or social media promotions were conducted. Access control ensured that only invited participants could respond; the survey link was not publicly posted.

### Participants

Eligible participants were health delegates from ERUs of the Red Cross or Red Crescent Movement who had participated in at least one humanitarian mission within the past 15 years. Inclusion criteria required previous deployment with a Health ERU and direct involvement in clinical care or coordination of medical humanitarian operations. Individuals were excluded if they had not been deployed with a health ERU or if their deployments were exclusively with non-health ERUs. Participation was entirely voluntary, with no financial or material incentives offered. Respondents could withdraw at any point before submission.

### Questionnaire

The questionnaire was developed based on a literature review and expert consultation with specialists in emergency medicine, palliative care, mental health, global health, and humanitarian operations. It was designed and administered using the LimeSurvey platform between 1 October and 31 December 2024.

The instrument assessed the following key domains:

- Demographic and professional characteristics (e.g., age, gender, roles, number of missions, region of deployment, and organisational affiliation — as many delegates are engaged with multiple organisations internationally).
- Perceived importance of including palliative care within humanitarian health response, measured using Likert-scale items.
- Perceived challenges to the provision of palliative care (e.g., resource constraints, ethical dilemmas, cultural factors).
- Perceived training needs (e.g., palliative care in humanitarian settings, pain and symptom management, communication and breaking bad news).

The draft survey underwent pilot testing with five experts (n = 5) representing emergency care, palliative care, health services research, global health, and mental health, as well as experienced ERU delegates. Feedback on clarity, structure, and logic was incorporated to improve face validity before dissemination.

### Data Collection and Measurement

The final survey comprised 35 items across six thematic sections: demographics; professional roles and experience; importance and readiness for palliative care; pain management and medication access; communication and breaking bad news; and training needs. The survey spanned approximately six pages online.

Items were presented in a fixed logical order, with adaptive branching for specific items (e.g., opioid management questions for prescribers only). Respondents could navigate backwards to review or change answers before submission. Response types included five-point Likert scales, dichotomous (Yes/No) questions, and open-text fields. Example items included: “How important do you believe it is to integrate palliative care into emergency response efforts?” (Not important at all - Extremely important), and “Have you received any training on how to manage opioids before any of your deployments?” (Yes/No). LimeSurvey automatically validated mandatory fields and performed complete checks before submission.

The complete questionnaire is provided in Supplementary Material 1. The survey was conducted and reported in accordance with the Checklist for Reporting Results of Internet E-Surveys (CHERRIES)^20^ (see Supplementary Material 2).

### Bias Control

To minimise potential biases, the survey was conducted anonymously to reduce social desirability bias. Standardised wording was used for all closed items to promote consistency.

### Handling of Missing Data

LimeSurvey automatically validated mandatory items to minimise missing responses. Only submissions with ≥50% completion were included in the analysis; entries with <50% completion were excluded. Time-stamp review confirmed data integrity, and only the first complete response was retained for any duplicate entries. No directly identifying personal data were collected. Duplicate responses were checked using survey metadata available within the platform.

### Analysis

We conducted a descriptive analysis of survey responses to summarise delegates’ perceptions, self-reported knowledge, preparedness, training needs, and perceived barriers to palliative care delivery in humanitarian missions. Categorical variables were presented as frequencies and percentages, while open-ended responses were analysed thematically to capture qualitative insights. No inferential tests were conducted because the objective was to describe current practices rather than test hypotheses.

Free-text responses (Q35) were analysed using rapid content analysis by two independent reviewers (HK & FM); discrepancies were resolved through discussion. Quantitative and qualitative data were analysed separately but interpreted together to provide complementary insights into the current state of palliative care readiness and the perceived needs of ERU delegates. Because the total number of invitees was unknown—due to onward circulation of the survey by focal points—response rates were calculated only for survey starts.

### Ethical Considerations

Before beginning the survey, participants viewed an online information page describing the study’s purpose, voluntary nature, anonymity, and data protection measures. Completion of the survey constituted informed consent. No identifiers were collected, and data were stored in accordance with the EU General Data Protection Regulation (GDPR) on password-protected institutional servers.

The study was approved by Brandenburg Medical School’s Ethics Committee (Reference 187032024-ANF) and endorsed by the institution’s Data Protection Officer, ensuring compliance with ethical and data protection standards.

## Results

### Participant characteristics

#### Demographic Data

Of 173 total submissions, 114 were included in the analysis (103 complete and 11 partial ≥50%), while 59 submissions (34.1%) were excluded for <50% completion. Among the 114 respondents included (Table 1), the largest age group was 45–54 years (35.1%, 40/114), followed by 55–64 years (28.9%, 33/114) and 34–44 years (16.7%, 19/114). Few were aged 25–34 (3.5%, 4/114), and none were under 25. Two-thirds of respondents were female (64.9%, 74/114), one-third were male (34.2%, 39/114), and one respondent did not indicate gender (0.9%, 1/114).

**Table 1.**
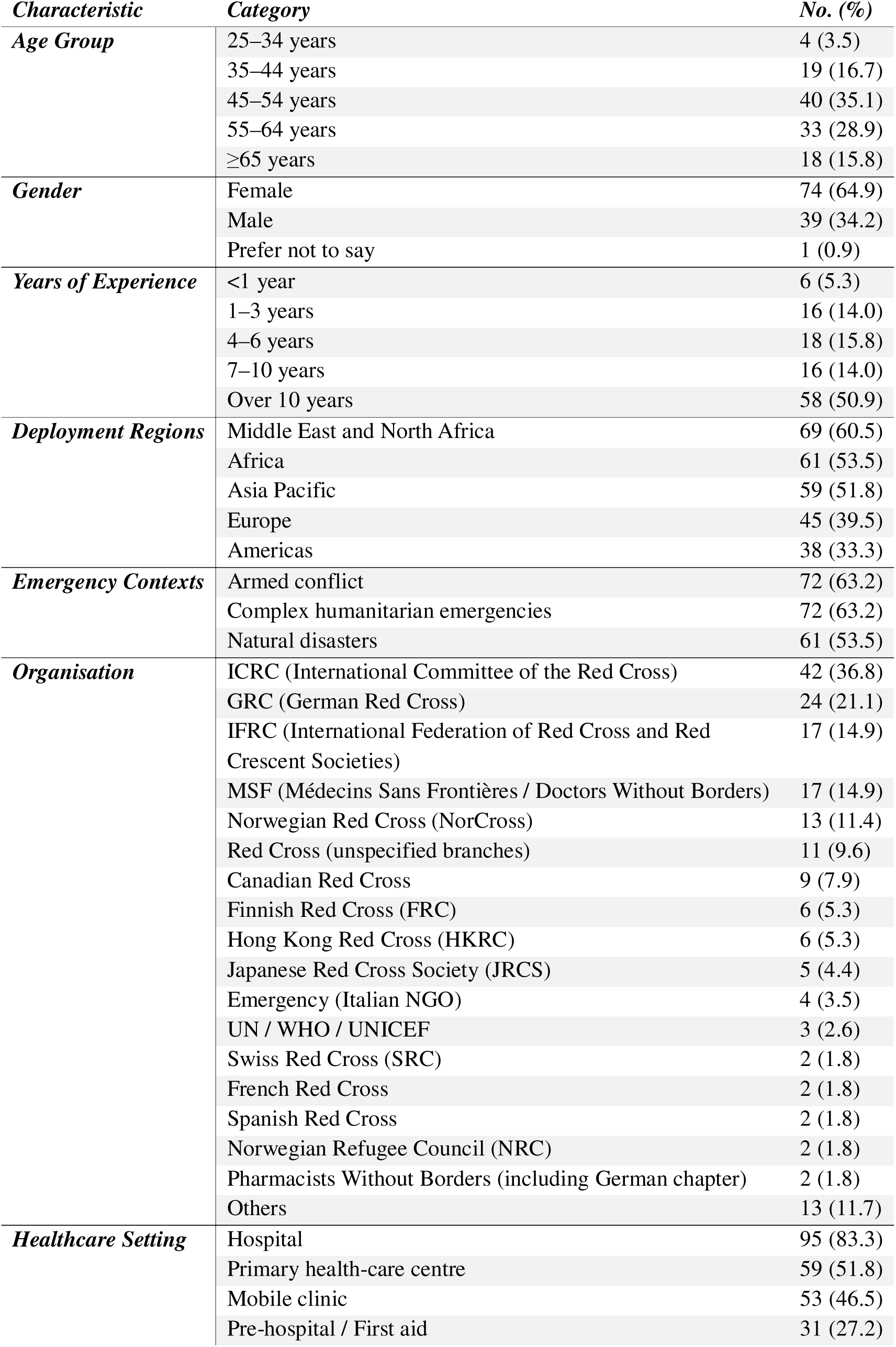
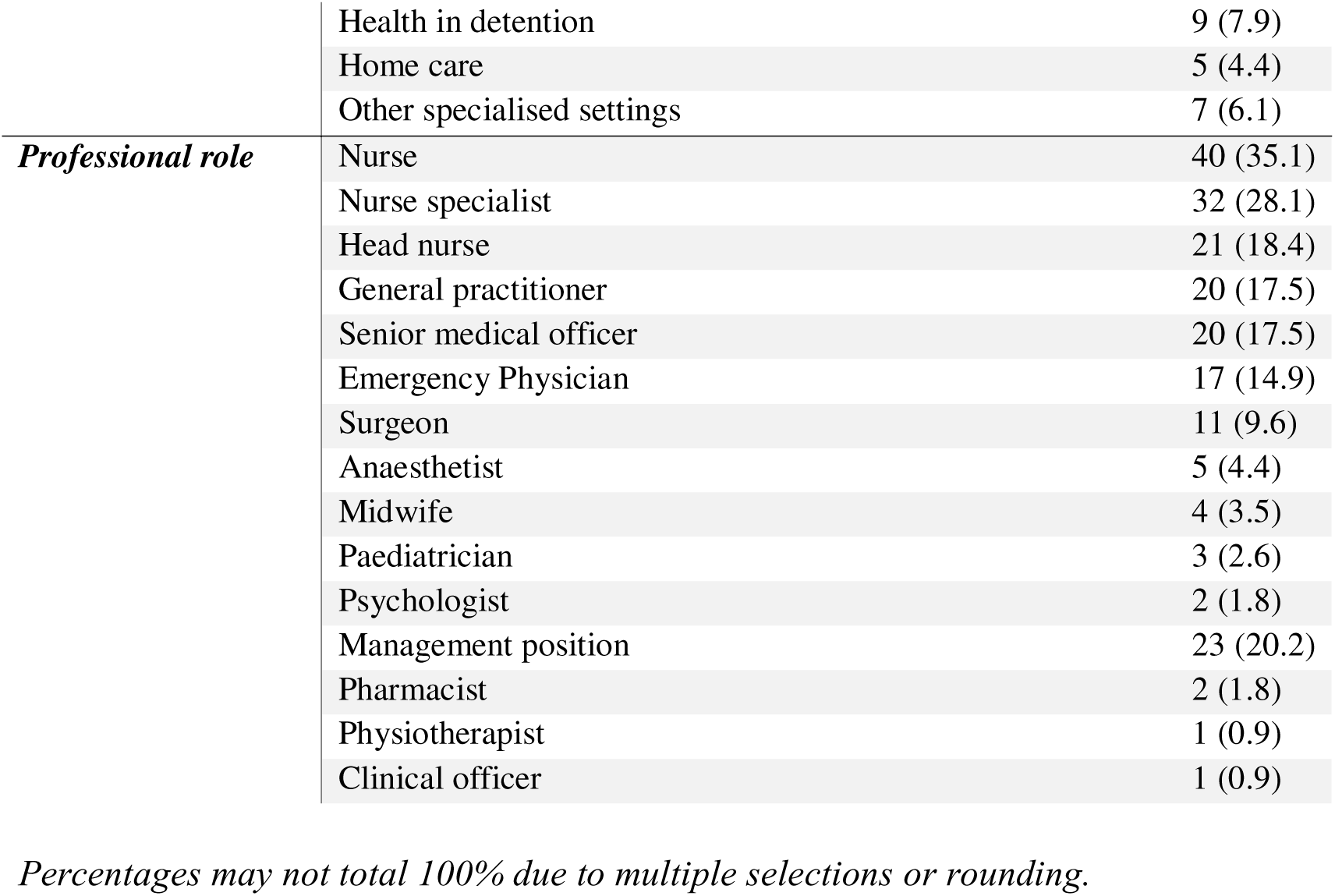
Participant Demographics, Affiliations, and Roles (n=114).

#### Professional Experience and Deployment Context

Half of the respondents (50.9%, 58/114) reported having over 10 years of humanitarian experience (Table 1). Smaller proportions were observed for 4–6 years (15.8%, 18/114), 7–10 years (14.0%, 16/114), 1–3 years (14.0%, 16/114), and less than one year of experience (5.3%, 6/114).

Participants had a broad range of deployment experience across regions, with the highest representation from the Middle East and North Africa (60.5%, 69/114), followed by Africa (53.5%, 61/114), Asia Pacific (51.8%, 59/114), the Americas (33.3%, 38/114), and Europe (39.5%, 45/114).

Regarding the nature of emergencies encountered, 63.2% (72/114) of respondents reported working in contexts of armed conflict, and an equal proportion (63.2%, 72/114) indicated experience in complex humanitarian emergencies. Additionally, 53.5% (61/114) were deployed in response to natural disasters.

#### Organisational Affiliation

Respondents represented a wide range of humanitarian organisations (Table 1). The largest group was from the International Committee of the Red Cross (36.8%, 42/114), followed by the German Red Cross (21.1%, 24/114), and both the International Federation of Red Cross and Red Crescent Societies and Médecins Sans Frontières (14.9%, 17/114 each). The Norwegian Red Cross was represented by 11.4% (13/114). Others came from various Red Cross and Red Crescent national societies—including those from Canada, Finland, Hong Kong, Japan, Switzerland, France, Spain, and the UK—as well as international organisations such as UN agencies, Emergency, and the Norwegian Refugee Council (NRC), reflecting broad sector representation.

#### Healthcare Settings and Roles

Most respondents (Table 1) reported deployment in hospital settings (83.3%, 95/114), followed by primary health care centres (51.8%, 59/114) and mobile clinics (46.5%, 53/114). Pre-hospital/first aid settings were also common (27.2%, 31/114), while fewer had experience in health in detention (7.9%, 9/114), home care (4.4%, 5/114), or other specialised settings (6.1%, 7/114), such as Ebola treatment centres and rescue ships.

In terms of professional roles, the largest groups were nurses (35.1%, 40/114) and nurse specialists (28.1%, 32/114), followed by head nurses (18.4%, 21/114), and general practitioners and senior medical officers (both 17.5%, 20/114). Additional roles included emergency physicians (14.9%, 17/114) and surgeons (9.6%, 11/114), as well as anaesthetists, midwives, paediatricians, psychologists, and management positions (20.2%, 23/114). Less frequently reported roles included pharmacists, physiotherapists, and clinical officers (each <2%).

#### Palliative Care during Deployments

When respondents were asked to define palliative care in humanitarian settings (Table 2), 86.0% (98/114) selected “all of the above,” indicating that palliative care includes physical, mental, social, and spiritual support. In addition, 29.8% (34/114) highlighted the importance of managing physical suffering, 26.3% (30/114) emphasised addressing mental and psychological issues, and 23.7% (27/114) viewed social support as a crucial component. Regarding the target of palliative care, nearly all respondents (90.4%; 103/114) believed that both patients and their relatives should receive care.

**Table 2.**
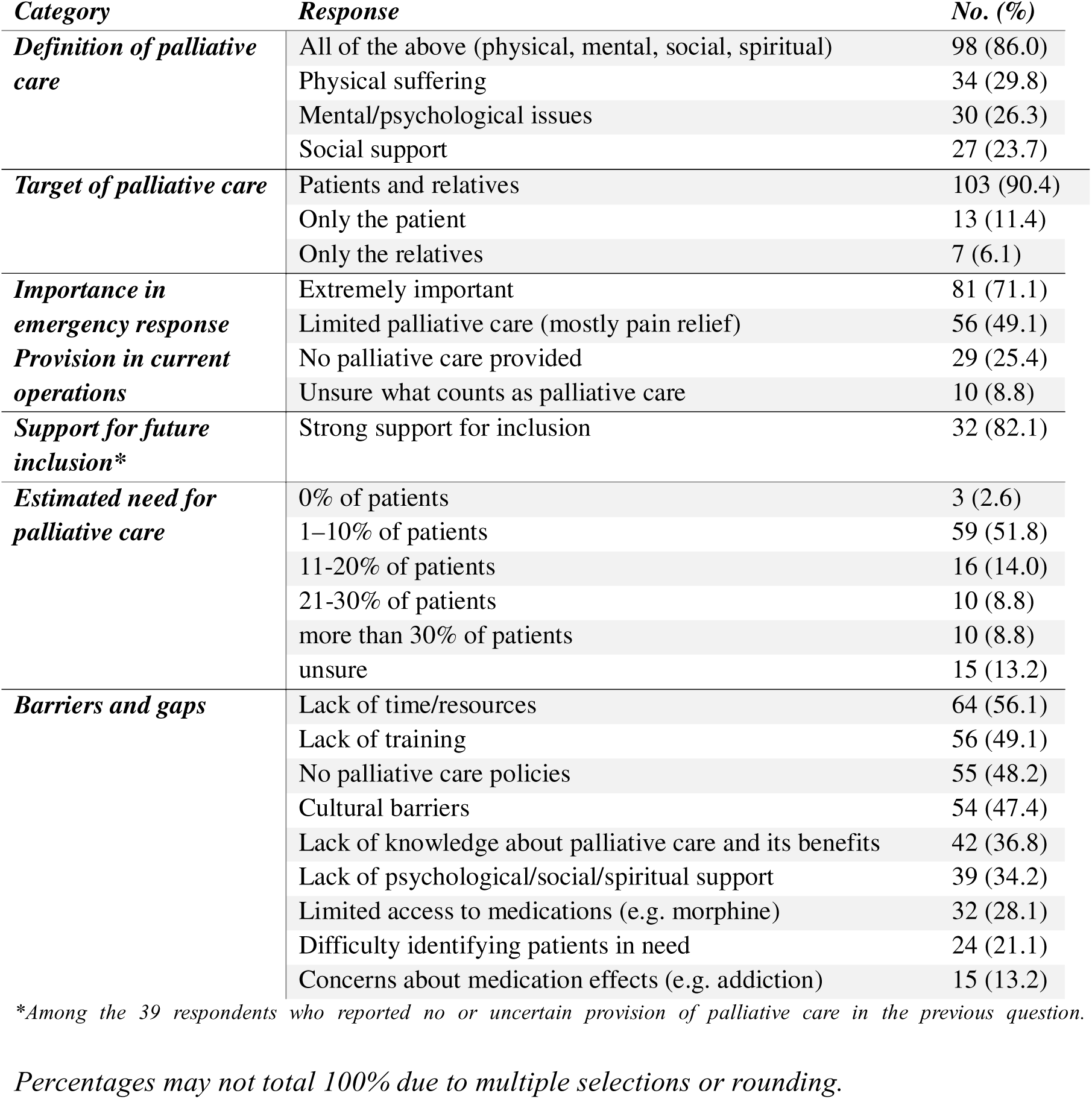
Palliative Care in Deployments / Perceptions and Provision of Palliative Care in Humanitarian Settings (n = 114).

Most respondents (71.1%, 81/114) considered it extremely important to integrate palliative care into emergency response efforts.

Around half of respondents (49.1%; 56/114) indicated that their organisations offered a limited range of palliative care services, primarily focusing on pain relief, and a quarter (25.4%; 29/114) reported that no palliative care was provided. Of those who did not offer palliative care (29/114) or were unsure what should be considered palliative care (10/114), 82.1% (32/39) expressed strong support for its inclusion in future efforts.

Regarding the estimated need for palliative care, over half of respondents (51.8%, 59/114) indicated that 1–10% of patients required palliative care, while 13.2% (15/114) were unsure.

When asked about the barriers or gaps encountered in providing palliative care during their deployments, respondents identified several key challenges (Figure 1). The most cited issues included insufficient time or resources (56.1%, 64/114), lack of palliative care training (49.1%, 56/114), and lack of knowledge about palliative care and its benefits (36.8%, 42/114). Other significant barriers included the absence of palliative care policies (48.2%, 55/114), difficulty identifying patients who may need palliative care (21.1%, 24/114), and limited access to essential medications, such as morphine or other opioids (28.1%, 32/114). Additional challenges included a lack of psychological, social, and spiritual support (34.2%, 39/114), concerns about the effects of medications (e.g., risk of addiction, 13.2%, 15/114), and cultural barriers to accepting palliative care (47.4%, 54/114).

**Figure 1:**
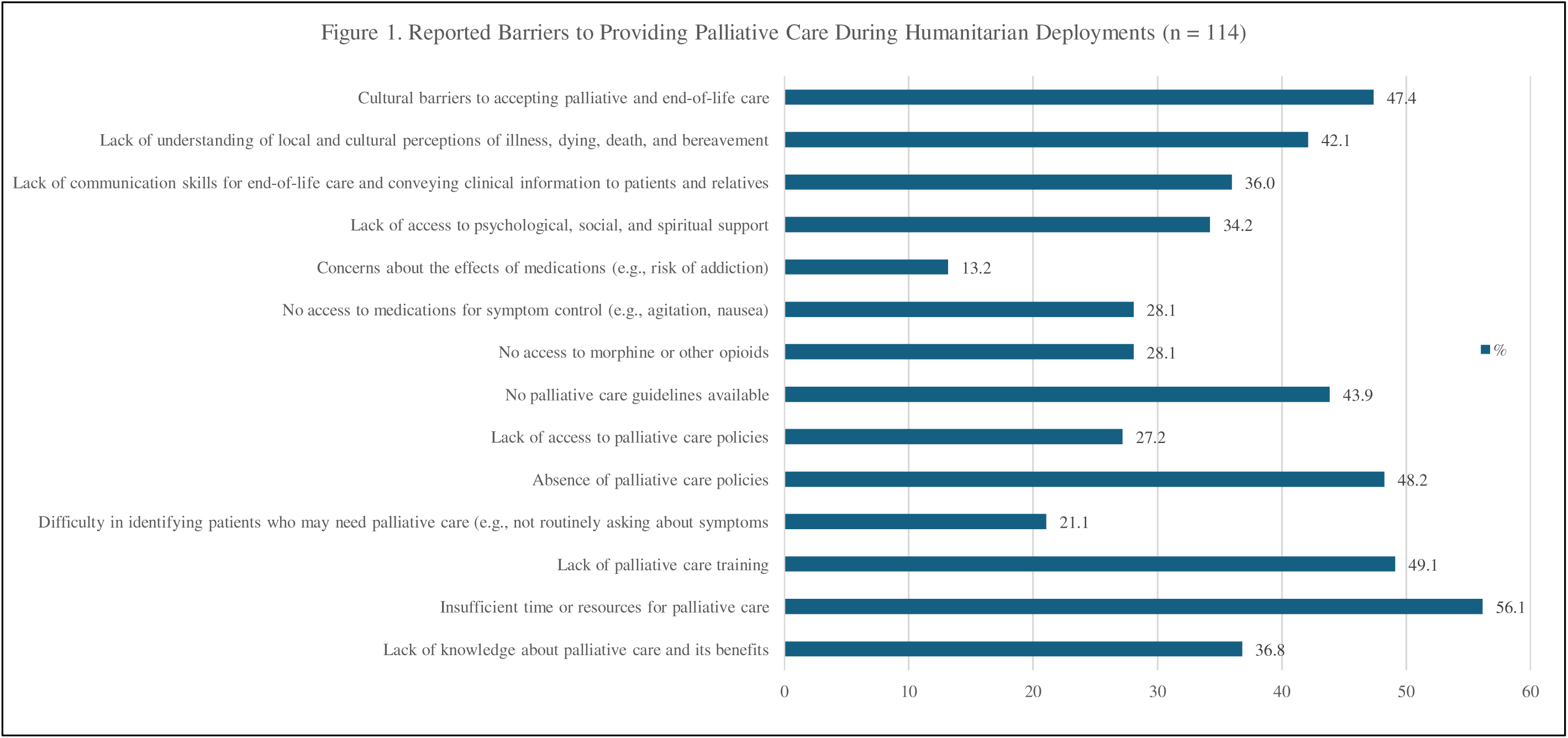
Reported Barriers to Providing Palliative Care During Humanitarian Deployments among Health ERU delegates (n = 114). Respondents could select multiple options.

#### Access to Opioids and Pain Management Challenges

During their deployments, two-thirds of respondents (64.9%; 74/114) reported access to weak or strong opioids (e.g., morphine, fentanyl, tramadol), while 28.9% (33/114) had no access, and 6.1% (7/114) indicated that this question did not apply to their situation (Table 3). For those without access to adequate pain management medications, 27.3% (9/33) were able to refer patients to a centre where they could receive such treatment, while 33.3% (11/33) could not, and 39.4% (13/33) were unsure.

**Table 3:** Pain Management during Deployments (N=114 unless otherwise noted).

| *Category* | *Response* | *No. (%)* |
| --- | --- | --- |
| *Access to opioids (morphine, fentanyl, tramadol)* | Had access | 74 (64.9) |
|  | Did not have access | 33 (28.9) |
|  | Not applicable | 7 (6.1) |
| *Referral options (among those without access, n = 33)* | Could refer patients to a centre | 9 (27.3) |
|  | Could not refer | 11 (33.3) |
|  | Not sure | 13 (39.4) |
| *Comfort prescribing opioids (among prescribers, n = 47)* | Comfortable | 40 (85.1) |
|  | Not comfortable | 1 (2.1) |
|  | Not sure | 6 (12.8) |
|  | Not prescribers (excluded from above) | 60 (of 107) (56.1) |
| *Medication administration routes (n = 107)* | Oral | 72 (67.3) |
|  | Intravenous (IV) | 67 (62.6) |
|  | Subcutaneous (SC) | 29 (27.1) |
|  | Enteral NG tube | 7 (6.5) |
|  | Rectal | 5 (4.7) |
|  | Transdermal | 2 (1.9) |
|  | Intramuscular (IM) | 35 (32.7) |
| *Challenges prescribing/administering meds (n = 107)* | Limited supply/stockouts | 58 (54.2) |
|  | Strict regulatory controls | 48 (44.9) |
|  | Concerns about misuse/addiction | 32 (29.9) |
|  | Lack of trained staff | 39 (36.4) |
|  | Absence of clear protocols/guidelines | 38 (35.5) |
|  | Cultural or religious concerns | 36 (33.6) |
| *Training on opioid management before deployment (n = 107)* | Received training | 51 (47.7) |
|  | Did not receive training | 56 (52.3) |

*Percentages may not total 100% due to multiple selections or rounding.*

Among respondents, 43.9% (47/107) identified as prescribers. Of these, 85.1% (40/47) felt comfortable prescribing opioids such as morphine or tramadol, 2.1% (1/47) did not, and 12.8% (6/47) were unsure.

Regarding medication administration, 67.3% (72/107) of respondents reported administering palliative care medications orally, 62.6% (67/107) intravenously, and 27.1% (29/107) subcutaneously. Other routes included enteral nasogastric tube (6.5%, 7/107), rectal (4.7%, 5/107), transdermal (1.9%, 2/107), and intramuscular (32.7%, 35/107).

Challenges in prescribing and administering palliative medications included limited supply or stockouts (54.2%, 58/107), strict regulatory controls on medications (44.9%, 48/107), and concerns about misuse or addiction (29.9%, 32/107). Other barriers were the lack of trained staff (36.4%, 39/107), absence of clear protocols or guidelines (35.5%, 38/107), and cultural or religious concerns (33.6%, 36/107).

Lastly, 47.7% (51/107) of respondents reported receiving training on how to manage opioids before their deployments, while 52.3% (56/107) did not.

### Communication and Delivering Bad News

Three-quarters of respondents (75.4%, 86/114) reported having to deliver bad news during deployments (Table 4), such as informing patients about serious medical conditions or notifying individuals of the death of a loved one. Only 18.4% (21/114) had not encountered such situations, and 6.1% (7/114) found this question not applicable to their experience. Two-thirds of respondents (62.8%; 54/86) did not use structured frameworks and approaches. Where used, these <u>included</u> SPIKES (12.8%, 11/86), the NURSE method (10.5%, 9/86), and other personal or professional approaches (e.g., Four Habits, empathic professional techniques). A small percentage had used the BREAKS (1.2%, 1/86), ABCDE (5.8%, 5/86), PEWTER (1.2%, 1/86), and REDMAP (3.5%, 3/86) frameworks. Regarding training, 27.9% (24/86) of those who had delivered bad news received training in communication techniques before their deployments, while 72.1% (62/86) did not.

**Table 4:** Delivering Bad News during Humanitarian Deployments — Frequency, Techniques, Challenges, and Training Needs (N=114 unless otherwise noted).

| *Category* | *Response* | *No. (%)* |
| --- | --- | --- |
| ***Had to deliver bad news during deployment*** | Yes | 86 (75.4) |
|  | No | 21 (18.4) |
|  | Not applicable | 7 (6.1) |
| ***Use of specific conversation techniques (Yes in previous question, n=86)*** | No | 54 (62.8) |
|  | SPIKES | 11 (12.8) |
|  | NURSE | 9 (10.5) |
|  | BREAKS | 1 (1.2) |
|  | ABCDE | 5 (5.8) |
|  | PEWTER | 1 (1.2) |
|  | REDMAP | 3 (3.5) |
| ***Challenges delivering bad news (n=107)*** | Language barriers | 86 (80.4) |
|  | Cultural/religious differences | 81 (75.7) |
|  | Emotional impact on healthcare providers | 27 (25.2) |
|  | Unclear prognosis | 32 (29.9) |
|  | Fear of patient/family reactions | 38 (35.5) |
| ***Training/support needed to improve delivery (n=107)*** | Cultural competency training | 77 (72.0) |
|  | Communication skills training | 54 (50.5) |
|  | Access to clear guidelines | 68 (63.6) |
|  | Psychological support for healthcare providers | 35 (32.7) |
|  | Peer support/mentoring | 45 (42.1) |

*Percentages may not total 100% due to multiple selections or rounding.*

Challenges encountered when delivering bad news included language barriers (80.4%, 86/107), cultural or religious differences (75.7%, 81/107), and the emotional impact on healthcare providers (25.2%, 27/107). Additional difficulties included unclear prognosis (29.9%, 32/107) and fear of patient or family reactions (35.5%, 38/107).

To improve their ability to deliver bad news effectively and compassionately, 72% (77/107) of respondents expressed the need for cultural competency training, while 50.5% (54/107) emphasised the importance of training on communication skills. Other desired support included access to clear guidelines (63.6%, 68/107), psychological support for healthcare providers (32.7%, 35/107), and peer support or mentoring (42.1%, 45/107).

### Training in Palliative Care for Humanitarian Contexts

Most respondents (82.7%, 86/104) had not received any prior training on palliative care in humanitarian settings (Table 5). However, 91.4% (96/105) of participants emphasised its importance. When asked about the preferred format for training, half of the respondents (50%;48/96) favoured blended learning, while 17.7% (17/96) preferred face-to-face training, and 14.6% (14/96) opted for online training.

**Table 5:** Training on Palliative Care in Humanitarian Settings (N=105).

| *Category* | *Response* | *N (%)* |
| --- | --- | --- |
| *Prior training on palliative care before deployment* | Yes | 18 (17.3) |
|  | No | 86 (82.7) |
| *Importance of specialised palliative care training* | Yes | 96 (91.4) |
|  | No | 9 (8.6) |
| *Preferred training format (N=96)* | Mixed (online and face-to-face) | 48 (50.0) |
|  | Face-to-face only | 17 (17.7) |
|  | Online only | 14 (14.6) |
|  | Other / Not specified | 17 (17.7) |
| *Preferred duration for online training (N=79)* | 2–4 hours | 37 (46.8) |
|  | 4–8 hours | 24 (30.4) |
|  | 8–20 hours | 11 (13.9) |
|  | Other / Not specified | 7 (8.9) |
| *Preferred duration for face-to-face training (N=77)* | One day | 31 (40.3) |
|  | Two days | 30 (39.0) |
|  | Three days | 16 (20.8) |
| *Face-to-face training should include practical components (N=82)* | Yes | 74 (90.2) |
|  | No | 8 (9.8) |
| *Basic palliative care knowledge included in ERU health training (N=105)* | Yes | 96 (91.4) |
|  | No | 9 (8.6) |
| *Important topics for training (N=105)* | Communication skills | 79 (75.2) |
|  | Assessment and management of pain | 81 (77.1) |
|  | Psychological and spiritual support | 74 (70.5) |
|  | Self-care and resilience for providers | 82 (78.1) |
|  | Ethical issues | 65 (61.9) |
|  | Cultural sensitivity | 69 (65.7) |
|  | Palliative care in epidemics | 65 (61.9) |

*Percentages may not total 100% due to multiple selections or rounding.*

Regarding the duration of online training, 46.8% (37/79) of respondents preferred sessions lasting 2-4 hours, 30.4% (24/79) preferred 4-8 hours, and 13.9% (11/79) preferred 8-20 hours. For face-to-face training, the largest proportion of respondents (40.3% (31/77)) suggested it should last for one day, followed by 39% (30/77) who preferred two days and 20.8% (16/77) who preferred three days. Additionally, 90.2% (74/82) believed that face-to-face training should include a practical component, underscoring the need for hands-on experience. A significant majority (91.4% (96/105)) also agreed that basic palliative care knowledge should be included in ERU Health training.

When asked about the topics to be included in palliative care training for humanitarian settings, key areas of interest were communication skills (75.2% (79/105)), assessment and management of pain (77.1% (81/105)), psychological and spiritual support for patients (70.5% (74/105)), and self-care and resilience for palliative care providers (78.1% (82/105)). Other priority topics included ethical issues (61.9% (65/105)), cultural sensitivity (65.7% (69/105)), and palliative care in epidemics of life-threatening infections (61.9% (65/105)).

### Open-ended feedback

Open-ended responses provided nuanced perspectives on how palliative care could be more effectively integrated into emergency deployments. Many respondents emphasised the need for field-ready standard operating procedures and checklists to embed palliative care into time-pressured environments. The majority highlighted communication and cultural safety as critical priorities, particularly for breaking bad news, working with interpreters, and engaging families in decision-making. Several respondents called for hands-on, humanitarian-specific training — such as simulation or on-mission refreshers — covering trauma-related suffering, psychological support, and context-appropriate symptom management. Others recommended embedding palliative care modules into ERU induction training, supported by practical tools such as dosing guides, communication prompts, and referral or escalation workflows.

A recurring theme was the importance of staff well-being and psychological support for both patients and responders. A few participants also identified systemic barriers to medicine access, noting that regulatory restrictions and stockouts often limit opioid availability. Where resources permit, respondents endorsed offline or tele-consultation tools for case discussions and guidance in the field.

## Discussion

### Principal findings

Our results indicate a substantial preparedness gap for palliative care among humanitarian health responders. Although respondents frequently encounter seriously ill and dying patients, only a minority reported prior formal training, and most emphasised an urgent need for practical, competency-based education. Communication emerged as a consistent challenge—particularly for breaking bad news across language barriers and navigating culturally specific expectations around truth-telling and decision-making. Medicine stockouts and restrictive policies impeded reliable access to opioids even where clinicians felt confident prescribing them. Identified training needs included: (1) culturally safe, high-stress communication; (2) pain and symptom management (opioid and non-opioid); (3) psychological and spiritual support; (4) cultural safety/sensitivity; and (5) staff well-being and self-care. Respondents favoured blended learning with strong practical components (i.e., simulation, scenarios, bedside coaching).

### Comparison with earlier research

These findings mirror and extend prior work showing that, despite WHO guidance, systematic integration of palliative care into humanitarian health operations remains the exception rather than the rule.^□,□,1□^ Earlier reviews and field studies describe similar barriers — limited training, absent or unclear protocols, and diffuse roles/responsibilities — and underscore that culturally attuned practice is essential but often under-resourced.^□,1□,1□^ Our data add EMT/ERU-specific operational details, including preferences for pre-deployment modules plus in-mission refreshers and the practical value of quick-reference tools for symptom control.

Access to essential controlled medicines remains a system-level bottleneck. Global analyses document wide disparities in opioid availability and highlight regulatory and supply-chain obstacles that are magnified in crises.^11^ Field case studies from humanitarian programmes in India and Bangladesh describe “opiophobia” *(the excessive fear of prescribing or using opioids for pain relief)*, complex licensing requirements, and recurrent stockouts that delay or prevent adequate analgesia, despite clear clinical indications.^12^ These observations align with normative guidance that calls for streamlined emergency provisions and coordinated action among ministries of health, narcotics control authorities, and humanitarian agencies.^1^□

### Implications for policy and practice

Translating guidance into routine practice requires action at three levels:

1. **People (competencies and culture):** Embed core palliative care competencies within standard ERU role descriptions and pre-deployment training, using simulation for communication and symptom-control scenarios typical of mass-casualty, displacement, or epidemic settings. Provide psychological first aid and structured peer support to mitigate moral distress.^□,1□,1□^
2. **Processes (Standard Operating Protocols (SOPs) and decision support):** Develop concise SOPs and quick-reference protocols mapped to EMT typology (Type 1/2/3) and resource levels, aligned with the WHO Blue Book and the WHO humanitarian palliative-care guide.^□,□^ Standardise a minimal palliative “kit” (e.g., oral morphine where legally feasible, adjuvants, antiemetics, antispasmodics, anxiolytics, non-pharmacologic measures). Introduce early goals-of-care triggers and structured family communication.
3. **Platforms (systems and partnerships):** Operationalise emergency controlled-medicine pathways and buffer stocks with IFRC/ICRC, WHO and national authorities; integrate palliative-care indicators into ERU monitoring and after-action reviews. Align with Sphere Health Standard 2.7 and IFRC Health & Care 2030 objectives to mainstream palliative care across preparedness, response, and recovery.^1^□,^1^□

### Strengths and limitations

A strength of this study is its focus on an extensive, multinational humanitarian network with standardised surge mechanisms, yielding insights directly actionable for EMT/ERU operations. However, the sampling frame intentionally targeted only Red Cross/Red Crescent (RCRC) delegates, limiting generalisability to other humanitarian actors and response models. In addition, the total number of invitees was unknown due to onward distribution, preventing calculation of a true denominator and response rate; as a result, the reported percentages reflect only the responding sample and may be affected by non-response and self-selection bias. Despite anonymisation, social desirability may also have influenced answers on sensitive topics (e.g., opioid use, end-of-life communication). Even so, the diversity of roles, geographies, and field experience among respondents supports the transferability and operational relevance of the findings. Lastly, while qualitative results were contextualised with quantitative findings, no methodological framing (such as triangulation or mixed-method integration framework) was applied.

### Recommendations for Action

Urgent efforts are needed to develop modular, flexible palliative-care training tailored to humanitarian responders — practical, culturally sensitive, and competency-based (communication; symptom control; psychological/spiritual support). These programmes could be delivered as a dedicated course, akin to the ICRC War Surgery Seminar, or embedded in the health ERU induction. Building on this foundation, implementation could proceed in phased steps:

Near-term (next 6–12 months): incorporate a palliative-care micro-curriculum into ERU induction (communication, analgesia basics, non-pharmacologic measures, cultural and ethical considerations); issue a one-page opioid access workflow tailored to national regulations where deployments are likely; pilot quick-reference SOPs in one ERU type with feedback loops.^□,□,11,12,1□^

Medium-term (12–24 months): formalise competency frameworks and assessment; pre-position essential medicines and adjuvants within ERU kits; launch the offline app with tele-mentoring capability; include palliative-care items and indicators in ERU readiness reviews and after-action reports; align with Sphere/IFRC frameworks to sustain adoption.^1^□,^1^□

Partnerships and platforms: major organisations (WHO, EAPC, WHPCA, APCA, and the Red Cross and Red Crescent Movement) should collaborate to systematically integrate palliative care across preparedness, response, and recovery^.□,1□,1□^

Digital and job aids: Develop an offline-capable, point-of-care tool hosting protocols/SOPs, dosing guides, communication prompts, and tele-consultation pathways for escalation in complex cases. In parallel, build context-adapted guidance, strengthen cultural competence training, and create psychological support systems for staff —recognizing the high emotional burden of providing palliative care in emergencies.^□,1□,1□^

### Future research

Research on the implementation of palliative care training programmes into ERU induction using CFIR 2.0 can identify determinants at the outer setting (regulatory environment, supply chains), inner setting (ERU culture, workflows), characteristics of individuals (confidence, skills), and characteristics of the intervention (SOPs, training, digital tools), guiding tailored strategies for scale-up and sustainment.^18^ Mixed-methods designs and realist evaluations can test how, for whom, and under what conditions training and decision support improve patient-and team-level outcomes across different crisis archetypes (conflict, natural disaster, epidemic).

### Conclusion

Palliative care is widely regarded by ERU delegates as integral to humanitarian response, yet provision remains inconsistent and often limited to analgesia. Respondents reported gaps in training, policies, communication skills, and access to essential medicines—barriers that persist despite high clinical need and a strong willingness among clinicians to provide care. Targeted, practical actions could close this implementation gap: integrate core palliative competencies into ERU induction and refresher training; adopt concise SOPs and decision aids mapped to ERU types; secure reliable access to essential controlled medicines through context-appropriate regulatory pathways; and strengthen culturally safe communication and psychological support for staff. Embedding these elements within routine preparedness, deployment, and after-action processes— aligned with WHO Emergency Medical Team standards and Red Cross and Red Crescent Movement frameworks—can normalise relief of suffering as a fundamental component of quality humanitarian healthcare.

## Declarations

HK, MK, MH and FM conceptualised and designed the study. Data collection and analysis were conducted by HK and FM. HK and FM drafted the manuscript, SM and MA contributed substantial revisions and interpretation of the findings. All authors reviewed and approved the final version of the manuscript.

## Consent for publication

Not applicable.

## Availability of data and materials

All data relevant to the study are included in the article or uploaded as supplementary material. Reasonable requests for additional information may be directed to the corresponding author.

## Data Availability

All data produced in the present study are available upon reasonable request to the authors

## Acknowledgements

The authors extend their sincere gratitude to all participants for generously sharing their time and insights. We are also grateful to the Red Cross and Red Crescent Movement for supporting the data collection of this study. Special thanks are due to the German Red Cross and to the Health Team at the International Federation of Red Cross and Red Crescent Societies in Geneva for their valuable support.

