## Supplementary material for "Palliative Care in Humanitarian Settings: An International Survey on Perceived Importance and Readiness among Health Emergency Response Unit Delegates": pdf

Survey

### Aim of the Survey

This survey is designed to assess and understand the need for enhancing palliative care services within both existing and new emergency response teams associated with the Red Cross and Red Crescent movement. We aim to explore the interest of Red Cross and Red Crescent health delegates in palliative care, taking into account their previous experiences and expectations of its importance in humanitarian settings.

The insights gathered from this survey will contribute to a detailed analysis of the current barriers and strengths in providing palliative care. Additionally, it will help identify the future training needs essential for supporting the delivery of palliative care in crisis situations.

### Context

Palliative care, as defined by the World Health Organization (WHO), is an approach that improves the quality of life of patients and their families who are facing life-threatening illness. This is achieved through the prevention and relief of suffering by means of early identification, and impeccable assessment and treatment of pain and other physical, psychosocial, and spiritual problems.

Your participation is vital in shaping the future of palliative care within the Red Cross and Red Crescent humanitarian response.

There are 35 questions in this survey.

### Section 1: Demographics

In this section, we will collect basic demographic information to better understand the background and context of the survey participants. This information will help us analyze responses in relation to various factors such as experience level and geographic location.

1. **What is your age? ***

Choose one of the following answers

Please choose **only one** of the following:

- 18-24
- 25-34
- 35-44
- 45-54
- 55-64
- 65 or older

2. **What is your gender? ***

Choose one of the following answers

Please choose **only one** of the following:

- Male
- Female
- Non-binary / Third gender
- Prefer not to say

3. **How many years of experience do you have working in emergency response or humanitarian settings? ***

Choose one of the following answers

Please choose **only one** of the following:

- • Less than 1 year
- • 1–3 years
- • 4–6 years
- • 7–10 years
- • More than 10 years

4. **Please specify the organization(s) you are currently deployed with or have been deployed with in the past 15 years.**

Please write your answer(s) here:

- 1
- 2
- 3
- 4
- 5

- 6
- 7
- 8
- 9
- 10

5. **Please select the region(s) where you are currently deployed or have been deployed in the past, and if possible, indicate the specific countries. ***

Comment only when you choose an answer.
Please choose all that apply and provide a comment:

- Americas
- Africa
- Middle East and North Africa MENA
- Europe
- Asia Pacific

6. **Please describe the type of emergency you are currently deployed in or have been deployed in: ***

Comment only when you choose an answer.
Please choose all that apply and provide a comment:

- Natural Disaster
- Armed Confilct
- Complex humanitarian emergency

Please provide additional details

7. **Please select the types of healthcare settings where you are currently deployed or have been deployed in the past from the list below: ***

Select all that apply
Please choose **all** that apply:

- Pre-hospital/ First Aid
- Primary health care center
- Mobile Clinic
- Hospital
- Health care in detention
- Home care
- Other:

8. **Please select the role(s) you have held during your current or previous deployments: ***

Select all that apply

Please choose **all** that apply:

- Nurse
- Nurse specialist
- Head nurse
- General physician
- Senior medical officer (Doctor)
- Paramedic
- Emergency Medicine Doctor (ER)
- Surgeon
- Anaesthetist
- Clinical officer
- Physiotherapist
- Psychologist
- Gynecologist
- Midwife
- Pediatrician
- Management
- Other:

**<u>Section 2: General Information about Palliative Care in Humanitarian Settings</u>**

In this section, we aim to gather insights into your understanding and experiences related to palliative care in humanitarian settings. Your responses will help us gauge current knowledge, assess the perceived importance of palliative care, and identify gaps and opportunities for development.

9. **What do you consider to be part of palliative care in humanitarian settings? ***

Select all that apply

Please choose **all** that apply:

- a. Management of physical suffering (e.g., pain and other symptoms)
- b. Addressing mental/psychological issues (e.g., grief, anxiety, depression)
- c. Providing social support (e.g., practical assistance, connection with others)
- d. Addressing spiritual issues (e.g., existential questions, spiritual distress)
- e. All of the above

**10. Who do you think is the target of palliative care in humanitarian settings? ***

Select all that apply

Please choose **all** that apply:

- a. Patient
- b. Relatives
- c. Both
- Other:

**11. How important do you believe it is to integrate palliative care into emergency response efforts? ***

Choose one of the following answers

Please choose **only one** of the following:

- • Extremely important
- • Moderately important
- • Slightly important
- • Not important at all

**<u>Section 3: Provision of Palliative Care and Perceived Barriers</u>**

This section focuses on understanding the current provision of palliative care in humanitarian settings and identifying the challenges faced. Your input will help highlight effective practices and areas needing improvement.

**12. Is/Was palliative care provided by your organization in your project setting during your deployment(s)? ***

Choose one of the following answers

Please choose **only one** of the following:

- Not sure about what should be considered palliative care in my setting
- Yes, we provide a comprehensive range of palliative care services (physical, mental, social, spiritual)
- Yes, but we provide only a limited number of services (e.g., mostly pain relief)
- No, we do not provide palliative care

Make a comment on your choice here:

*In the Comment: Please elaborate on the types of services provided by your organization/project (physical, mental, social, spiritual, or other):*

13. **Do you think palliative care should be provided in your project? ***

*Only answer this question if the following conditions are met:*
*Answer was 'No, we do not provide palliative care' or 'Not sure about what should be considered palliative care in my setting' at question ' [G03Q08]' (12. Is/Was palliative care provided by your organization in your project setting during your deployment(s)?)*

Choose one of the following answers
Please choose **only one** of the following:

- Yes
- No

Make a comment on your choice here:
Please describe why you think this is the reason

14. **What percentage of patients attending the healthcare facility you are currently working in or have worked in during your deployment(s) do you estimate need palliative care? ***

Choose one of the following answers
Please choose **only one** of the following:

- 0%
- 1-10%
- 11–20%
- 21–30%
- More than 30%
- Unsure
- Other

15. **During your deployment(s), what gaps or barriers have you encountered in providing palliative care? ***

Select all that apply
Please choose **all** that apply:

- Lack of knowledge about palliative care and its benefits
- Insufficient time or resources for palliative care
- Lack of palliative care training
- Difficulty in identifying patients who may need palliative care (e.g., not routinely asking about symptoms)
- Absence of palliative care policies
- Lack of access to palliative care policies
- No palliative care guidelines available
- No access to morphine or other opioids
- No access to medications for symptom control (e.g., agitation, nausea)
- Concerns about the effects of medications (e.g., risk of addiction)
- Lack of access to psychological, social, and spiritual support

- Lack of communication skills for end-of-life care and conveying clinical information to patients and relatives
- Lack of understanding of local and cultural perceptions of illness, dying, death, and bereavement
- Cultural barriers to accepting palliative and end-of-life care
- Other:

**Section 4: Clinical Practice – Access to Medication, Prescribing, Administration, and Breaking Bad News**

This section addresses the availability of medications, the confidence in prescribing and administering them, as well as the challenges and techniques involved in delivering difficult news in palliative care settings.

16. **During your deployment(s), Do you have access to weak or strong opioids (e.g., Morphine, Fentanyl, Tramadol, codeine etc.)? ***

Please choose **only one** of the following:

- Yes
- No

17. **If you do not have access to adequate medication for the pain management of a palliative care patient (e.g., Morphine, Tramadol, or other opioids), is/was there a center you can refer the patient to for that service? ***

*Only answer this question if the following conditions are met:*
*Answer was 'No' at question ' [G05Q13]' (16. During your deployment(s), Do you have access to weak or strong opioids (e.g., Morphine, Fentanyl, Tramadol, codeine etc.)? )*

Choose one of the following answers

Please choose **only one** of the following:

- Yes
- No
- Unsure / I don't know

Make a comment on your choice here:

If you answered "Yes" and know the distance to the nearest referral center (in km), please specify it:

18. **If you are a prescriber, are you comfortable with prescribing medications (Morphine, Tramadol, or other opioids) in a palliative care context? ***

Choose one of the following answers

Please choose **only one** of the following:

- I am not a prescriber
- Yes
- No
- Unsure

Make a comment on your choice here:
If No or Unsure, please explain the reason:

19. **How is/was medication for palliative care administered during your deployment(s)? ***

Select all that apply
Please choose **all** that apply:
- Oral administration
- Intravenous
- Enteral via NG tube
- Subcutaneous
- Transdermal
- Rectal
- Intramuscular
- Other:

20. **During your deployment(s), what challenges do you encounter in prescribing and administering these medications? ***

Select all that apply
Please choose **all** that apply:
- Strict regulatory controls on medications (e.g., opioids)
- Lack of trained staff to prescribe/administer medications
- Limited supply or stockouts of medications
- Concerns about potential misuse or addiction
- Lack of protocols or guidelines for prescribing and administration
- Cultural or religious concerns about certain medications
- Other:

21. **Have you received any training on how to manage opioids before any of your deployments? ***

Choose one of the following answers
Please choose **only one** of the following:
- Yes
- No

Make a comment on your choice here:
• If Yes, please explain more:

**22. Have you ever been in a situation during your deployment(s) where you had to deliver bad news (e.g., informing a patient about a serious medical condition, notifying someone of a loved one's death)? ***

Please choose **only one** of the following:

- Yes
- No

**23. Have you utilized specific conversation techniques for delivering/breaking bad news to patients and their families? ***

*Only answer this question if the following conditions are met:*
*Answer was 'Yes' at question ' [G03Q18]' (22. Have you ever been in a situation during your deployment(s) where you had to deliver bad news (e.g., informing a patient about a serious medical condition, notifying someone of a loved one's death)?)*

Select all that apply

Please choose **all** that apply:

- Yes, SPIKES model
- Yes, BREAKS Protocol
- Yes, ABCDE Model
- Yes, PEWTER Approach
- Yes, NURSE Technique
- Yes, REDMAP Framework
- No
- Other:

**24. Have you received any training on these techniques before any of your deployments? ***

*Only answer this question if the following conditions are met:*
*Answer was 'Yes' at question ' [G03Q18]' (22. Have you ever been in a situation during your deployment(s) where you had to deliver bad news (e.g., informing a patient about a serious medical condition, notifying someone of a loved one's death)?)*

Choose one of the following answers

Please choose **only one** of the following:

- Yes
- No

Make a comment on your choice here:

If Yes, please explain more in the comment

**25. What challenges do you encounter when delivering bad news during your deployments? ***

Select all that apply

Please choose **all** that apply:

- Language barriers

- Cultural or religious differences in handling bad news
- Lack of training on communication skills
- Emotional impact on the healthcare provider
- Unclear prognosis or medical uncertainty
- Fear of patient or family reactions
- Other:

26. **What support or training would help you improve your ability to break bad news effectively and compassionately? ***

Select all that apply

Please choose **all** that apply:

- Training on communication skills
- Cultural competency training
- Access to clear guidelines or protocols
- Psychological support for healthcare providers
- Peer support or mentoring
- Other:

**<u>Section 5: Perceived Need for Training</u>**

This section aims to gather information on the perceived need for training in palliative care among healthcare professionals working in humanitarian settings.

27. **Have you received any training on Palliative Care in Humanitarian Settings before any of your deployments? ***

Choose one of the following answers

Please choose **only one** of the following:

- No
- Yes

Make a comment on your choice here:

If Yes, please specify the type of training and which deployment(s) it was before

28. **Do you see a need for specialized training in palliative care support tailored to humanitarian settings? ***

Choose one of the following answers

Please choose **only one** of the following:

- Yes
- No

Make a comment on your choice here:

Please explain your reasoning:

**29. What format would you prefer for palliative care training, and why? ***

*Only answer this question if the following conditions are met:*
*Answer was 'Yes' at question ' [G01Q25]' (28. Do you see a need for specialized training in palliative care support tailored to humanitarian settings? )*

Choose one of the following answers
Please choose **only one** of the following:

- Online
- Face-to-Face
- Either
- Mixed (Online and Face-to-Face)

Make a comment on your choice here:

**30. For online training, how long do you think the training should last? ***

*Only answer this question if the following conditions are met:*
*Answer was 'Online' or 'Either' or 'Mixed (Online and Face-to-Face)' at question ' [G01Q26]' (29. What format would you prefer for palliative care training, and why? )*

Choose one of the following answers
Please choose **only one** of the following:

- 2-4 hours
- 4-8 hours
- 8-20 hours
- More than 20 hours

Make a comment on your choice here:

**31. For Face-to-Face training, how long do you think the training should last? ***

Only answer this question if the following conditions are met:
Answer was 'Face-to-Face' *or* 'Either' *or* 'Mixed (Online and Face-to-Face)' at question ' [G01Q26]' (29. What format would you prefer for palliative care training, and why? )

Choose one of the following answers
Please choose **only one** of the following:

- 1 Day
- 2 Days
- 3 Days
- Longer than 3 Days

Make a comment on your choice here:

32. **Do you think Face-to-Face training should include a practical component? ***

*Only answer this question if the following conditions are met:*
*Answer was 'Face-to-Face' or 'Either' or 'Mixed (Online and Face-to-Face)' at question '[G01Q26]' (29. What format would you prefer for palliative care training, and why? )*

Choose one of the following answers

Please choose **only one** of the following:

- Yes
- No

Make a comment on your choice here:

33. **Do you believe that basic knowledge of palliative care should be included in the Emergency Response Unit (ERU) Health training? ***

Choose one of the following answers

Please choose **only one** of the following:

- Yes
- No

Make a comment on your choice here:

34. **What topics should be included in palliative care training for humanitarian settings? ***

Select all that apply

Please choose **all** that apply:

- Enrollment Criteria for Palliative Care
- Communication Skills (e.g., breaking bad news, active listening, ...)
- Nursing Care in Advanced Illness
- Assessment and Management of Pain (incl. opioid use)
- Assessment and Management of Other Symptoms (e.g., agitation, nausea, breathlessness)
- Mental Health in Palliative Care (e.g., anxiety, depression)
- Psychological and spiritual support for patients
- Psychosocial, Spiritual, and Bereavement Support for Families
- Self-Care and Resilience for Palliative Care Providers
- Neonatal and Pediatric Palliative Care
- End-of-Life Care Planning / decision-making
- Palliative Care in Epidemics of Life-Threatening Infections
- Community Engagement Regarding Palliative Care
- Ethical issues in palliative care
- Cultural sensitivity in palliative care

35. **Do you have any additional comments, suggestions, or concerns related to palliative care in humanitarian settings that were not addressed in the survey?**

Please write your answer here:

**Thank You for your participation!**

Your feedback is invaluable in shaping the future of palliative care within humanitarian settings. Together, we can enhance the quality of care provided to those in need, ensuring that every patient receives compassionate and comprehensive support during times of crisis.

Your contribution will help us make a meaningful impact. We appreciate your time and effort in providing us with your insights and experiences.

If you are interested in the topic of palliative care in humanitarian settings, please feel free to email me to express your interest. This will help us coordinate any future work in this area.

**Thank you for completing this survey.**
