## Supplementary material for "Palliative Care in Humanitarian Settings: An International Survey on Perceived Importance and Readiness among Health Emergency Response Unit Delegates": pdf

| Checklist item | Explanation | Description in this study | Where reported |
|---|---|---|---|
| ***Study design*** | Target population, sampling frame | Cross-sectional, web-based survey of Red Cross/Red Crescent Health ERU delegates with deployment experience (2010–2024). | Methods – Study design & purpose; Participants |
| ***IRB / Ethics*** | IRB approval noted | Brandenburg Medical School Ethics Committee (Ref. 187032024-ANF); Data Protection Officer (DPO) endorsement; anonymous participation. | Ethical Considerations |
| ***Informed consent*** | Consent process | Online information page; voluntary; proceeding implied consent; anonymity and data use explained. | Ethical Considerations |
| ***Data protection*** | Protection of personal data | No identifiers collected; GDPR-compliant storage on password-protected institutional servers. | Ethical Considerations |
| ***Development & testing*** | Survey development/usability testing | LimeSurvey instrument; piloted with 5 experts (emergency, palliative, HSR) + experienced delegates; wording/logic refined. | Data Collection and Measurement |
| ***Open vs closed*** | Access to the survey | Closed survey; secure link shared via internal IFRC/ICRC lists and National Society focal points; not publicly posted. | Methods – Setting |
| ***Contact mode*** | How invitees were contacted | Email invitations through IFRC/ICRC clinical delegate pools and National Societies. | Methods – Setting |
| ***Advertising*** | Announcements/ads | None; no public advertisement or social media. | Methods – Setting |
| ***Web / E-mail*** | Mode of survey | Web survey hosted on LimeSurvey (secure link). | Methods – Setting |

| | | | |
|---|---|---|---|
| ***Context*** | Host website context | Not posted on a public site; access via invitation link only. | Methods – Setting |
| ***Mandatory/voluntary*** | Requirement to participate | Voluntary; no incentives, withdrawal possible before submission. | Methods – Participants |
| ***Incentives*** | Incentives offered | None. | Methods – Participants |
| ***Timeframe*** | Field period | 1 Oct–31 Dec 2024. | Methods – Setting |
| ***Reminder(s)*** | Follow-ups during fieldwork | One reminder email mid-field period. | Methods – Setting |
| ***Randomization of items*** | Item order/alternation | No randomization; fixed logical order. | Methods – Data Collection & Measurement |
| ***Adaptive questioning*** | Branching/skip logic | Yes; conditional items (e.g., training and prescriber items based on prior answers). | Methods – Data Collection & Measurement |
| ***Number of items*** | Items per page | 35 standardized items across thematic sections. | Methods – Data Collection & Measurement |
| ***Number of pages*** | Screens/pages | ~6 online pages across six thematic sections. | Methods – Data Collection & Measurement |
| ***Completeness check*** | Validation prior to submit | LimeSurvey validation for mandatory fields; inclusion threshold ≥50% completion. | Methods – Data collection & measurement; Handling of missing data |
| ***Review step*** | Ability to change answers | Back navigation permitted prior to submission. | Methods – Data Collection & Measurement |

| | | | |
|---|---|---|---|
| ***Unique visitor definition*** | Prevent/handle duplicates | Single universal link; duplicate entries screened by submission timestamps/response patterns; first complete retained. | Methods – Setting |
| ***View rate*** | Unique survey visitors / site visitors | Not applicable (closed survey; denominator unknown due to onward distribution). | Methods – Analysis |
| ***Participation rate*** | Agreed / first page visitors | Reported among survey starts; 173 submissions total. | Results – Participants |
| ***Completion rate*** | Finished / agreed | 114/173 included (103 complete + 11 partial ≥50%); 59/173 excluded <50%. | Results – Participants |
| ***Cookies used*** | Cookie policy | No cookies used or stored. | Methods - Handling of missing data |
| ***IP check*** | IP storage/use | No IP addresses collected or stored. | Methods - Handling of missing data |
| ***Log file analysis*** | Other duplicate checks | Manual screening of submission timestamps, no additional log analyses. | Methods – Analysis |
| ***Registration*** | Login/registration | No login; access restricted via invitation links to ERU pools. | Methods – Setting |
| ***Handling incomplete*** | Inclusion/exclusion rules | ≥50% completion included in descriptive summaries; <50% excluded. | Methods – Handling Missing Data |
| ***Atypical timestamps*** | Very short completion handling | Not used as exclusion criterion; manual integrity checks only. | Methods – Handling of missing data |
| ***Statistical correction*** | Weighting/propensity scores | None; descriptive statistics; free-text rapid content analysis (two reviewers; consensus). | Methods – Analysis |

| | | | |
|---|---|---|---|
| ***Checklist citation*** | Reference to CHERRIES | Cited Eysenbach 2004 (JMIR). | Methods – Data collection & measurement; References (#20) |
| ***Questionnaire availability*** | Access to instrument | Full questionnaire in Supplementary Material 1 (PDF). | Methods – Data collection & measurement |
